# SARS-CoV-2 variants of concern exhibit reduced sensitivity to live-virus neutralization in sera from CoronaVac vaccinees and naturally infected COVID-19 patients

**DOI:** 10.1101/2021.07.10.21260232

**Authors:** Vimvara Vacharathit, Pakorn Aiewsakun, Suwimon Manopwisedjaroen, Chanya Srisaowakarn, Thanida Laopanupong, Natali Ludowyke, Angsana Phuphuakrat, Chavachol Setthaudom, Supanuch Ekronarongchai, Sirawat Srichatrapimuk, Pattama Wongsirisin, Suleeporn Sangrajrang, Thanarath Imsuwansri, Suppachok Kirdlarp, Sureeporn Nualkaew, Insee Sensorn, Waritta Sawaengdee, Nuanjun Wichukchinda, Somnuek Sungkanuparph, Wasun Chantratita, Mongkol Kunakorn, Jinda Rojanamatin, Suradej Hongeng, Arunee Thitithanyanont

## Abstract

Recent surges in SARS-CoV-2 variants of concern (VOCs) call for the need to evaluate levels of vaccine-and infection-induced SARS-CoV-2 neutralizing antibodies (NAbs). CoronaVac (Sinovac Biotech, Beijing, China) is currently being used for mass vaccination in Thailand as well as other low-income countries. Three VOCs currently circulating within Thailand include the B.1.1.7 (Alpha), B.1.351 (Beta), and B.1.617.2 (Delta) strains. We assessed NAb potency against the prototypic strain containing the original spike sequence (WT) compared to that against the 3 VOCs using sera derived from a cohort of healthcare workers who received a full 2-dose regimen of CoronaVac. Sera from two other cohorts consisting of COVID-19 patients who had been hospitalized in 2020 and 2021 were evaluated for comparison. We found that, despite equally robust production of S1-RBD-binding IgG and 100% seropositivity, sera from both CoronaVac vaccinees and naturally infected individuals had significantly reduced neutralizing capacity against all 3 VOCs compared to WT. Strikingly, NAb titers against Alpha and Beta were comparable, but Delta appears to be significantly more refractory to NAbs in all groups. Our results may help inform on CoronaVac NAb-inducing capacity, which is a proxy for vaccine efficacy, in the context of the WT strain and 3 VOCs. Our results also have critical implications for public health decision-makers who may need to maintain efficient mitigation strategies amid a potentially high risk for infection with VOCs even in those who have been previously infected.

## Introduction

The rapid emergence and spread of SARS-CoV-2 variants of concern (VOCs) harboring diverse mutations within the vaccine-targeted spike (S) protein have raised concerns over their immune and vaccine evasion potential. The inactivated whole-virus vaccine CoronaVac (Sinovac Biotech, Beijing, China) has been government-approved for emergency use in mass vaccination programs in Thailand and is widely available in many low-income countries. Results from a phase 1/2 clinical trial for CoronaVac were recently published (1). A large observational study in Chile further estimated that full immunization with CoronaVac had adjusted vaccine effectiveness of 65.9% for the prevention of COVID-19, 87.5% for hospitalization, 90.3% for ICU admission, and 86.3% for death (2). VOCs currently circulating in Thailand include B.1.1.7 (Alpha), B.1.351 (Beta), and B.1.617.2 (Delta). To assess the impact of SARS-CoV-2 variants on vaccine- and infection-induced antibodies, we evaluated S1-RBD-binding IgG levels and neutralizing antibody (NAb) titers against the SARS-CoV-2 prototypic vaccine strain (WT), the Alpha, Beta, and Delta strains in sera from healthcare workers who had received 2 doses of the CoronaVac; these results were compared to neutralization in sera from unvaccinated, naturally infected COVID-19 patients who had been hospitalized from March 2020 to May 2020 (denoted ‘Natural Infection 2020’) or in April 2021 to May 2021 (denoted ‘Natural Infection 2021’).

## Methods

### Cohort

Patients were confirmed to be infected by RT-PCR on nasopharyngeal and throat swab specimens through amplification of SARS-CoV-2 ORF1AB and N target gene fragments (Sansure Biotech Inc, Changsha, PR China). Details regarding patient demographics can be found in **Table 1**.

**Table 1.** Cohort demographics of CoronaVac vaccinees as well as naturally infected patients hospitalized in 2020 and 2021 who were included in the study.

| Characteristic |  | CoronaVac | Natural infection 2020 | Natural infection 2021 | P-value ‡ |  |
| --- | --- | --- | --- | --- | --- | --- |
|  |  |  |  |  | Log<sub>10</sub>(IgG titer) predictor | Log<sub>10</sub>(NAb titer) predictor |
| Sample – no. |  | 60 | 30 | 30 |  | <0.001 |
| Age – median (IQR*) |  | 35 (29.75–44.50) | 44.80 (36.80–55.85) | 46.45 (34.45–53.00) | 0.414 | 0.675 |
| Female – no. (%) |  | 51 (85%) | 11 (37%) | 11 (37%) | 0.682 | 0.013 |
| Days from latest vaccine dose or from illness onset – median (IQR*) |  | 15 (15–15.25) | 17.50 (15.25–21) | 17.50 (15–21.75) | <0.001 |  |
| IgG titer – median (IQR*) |  | 1508.4 (698.8–2320.1) | 2706.9 (1013.7–9139.5) | 3777.2 (1500.8–5068.8) |  | <0.001 |
| Neutralizing antibody titer<sup>†</sup> – median (IQR*) | WT | 640 (320–1280) | 2560 (800–10240) | 1280 (640–2635) |  |  |
|  | Alpha | 40 (17.5–40) | 320 (80–640) | 640 (320–1280) |  |  |
|  | Beta | 20 (10–40) | 320 (100–1120) | 480 (160–640) |  |  |
|  | Delta | 10 (10–40) | 80 (25–160) | 160 (50–320) |  |  |

\*IQR denotes inter-quartile range
†Neutralizing antibody titers below the limit of detection (<10) were assigned a value of 10
‡See the models in **Methods**

### Virus variants and culture

The wildtype SARS-CoV-2 virus (SARS-CoV-2/human/THA/LJ07_P3/2020), was isolated from a nasopharyngeal swab sample from an RT-PCR confirmed COVID-19 patient provided by Bamrasnaradura Infectious Diseases Institute, Nonthaburi, Thailand. African green monkey (Cercopithecus aethiops) kidney epithelial cells (Vero cells) (ATCC CCL-81) were used for virus isolation and a Vero cell derivative (Vero E6 cells) (ATCC CRL-1586) was used for virus propagation.

The B.1.1.7 (SARS-CoV-2/human/THA/NH657_P3/2021) and B.1.617.2 (SARS-CoV-2/human/THA/OTV007_P3/2021) variants were isolated from nasopharyngeal swab samples from RT-PCR confirmed COVID-19 patients provided by Ramathibodi Chakri Naruebodindra Hospital (Chakri Naruebodindra Medical Institute), Samut Prakan, Thailand. The B.1.351 variant (SARS-CoV-2/human/THA/NH088_P3/2021) was isolated from a nasopharyngeal swab sample from an RT-PCR confirmed COVID-19 patient provided by the Division of Genomic Medicine and Innovation Support, Department of Medical Sciences, Ministry of Public Health, Nonthaburi, Thailand. For B.1.1.7, B.1.617.2 and B.1.351, Vero E6 cells were used for both virus isolation and propagation.

Vero cells were cultured in minimum essential medium (MEM) (Gibco, Detroit, MI, USA) and Vero E6 were cultured in Dulbecco’s modified Eagle’s medium (DMEM) (Gibco). Virus isolation and propagation were conducted in a certified BSL3 facility at the Microbiology Department, Faculty of Science, Mahidol University, Bangkok, Thailand.

### Sequencing

Nucleic acid was extracted from 200 µl culture supernatant using the GenTiTM32 Automatic Extraction System (Advanced Viral DNA/RNA Extraction Kit) according to the manufacturer’s instructions, followed by library preparation using the ARTIC SARS-CoV-2 sequencing protocol (https://dx.doi.org/10.17504/protocols.io.bgxjjxkn). Prepared libraries were sequenced (paired-end) with single indexing on a MiSeq (Illumina) sequencer at the Center of Medical Genomics, Ramathibodi Hospital, Thailand.

The sequence of a reference SARS-CoV-2 (NCBI Reference Sequence: NC_045512.2) was used as a query sequence to pull SARS-CoV-2 reads from the Next Generation Sequencing read datasets by BLASTn. The BLASTn results were then used to construct alignments of the reads. After manual curation, the consensus sequences were determined according to the top 80% most frequent nucleotide residues at each position.

### Live-virus microneutralization

Sera were heat inactivated at 56°C for 30 min then two-fold serially diluted starting from 1:10. Equal volumes of SARS-CoV-2 were spiked into the serial dilutions at an infectious dose of 100 TCID_50_ (50% tissue culture infectious dose) and incubated for 1h at 37°C. Vero E6 cells (ATCC USA) were pre-seeded in Dulbecco’s modified Eagle medium (DMEM) supplemented with 2% fetal bovine serum (FBS), 100 U/mL penicillin, and 0.1 mg/mL of streptomycin. 100 µL of the virus-serum mixtures at different dilutions were added to 1×10_4_ pre-seeded Vero E6 cell monolayers in duplicate on a 96-well microtiter plate, then incubated for 2 days at 37°C and 5% CO_2_. The last two columns contained the virus control, cell control, and virus back-titration. Medium was discarded and cells were fixed and permeabilized with ice-cold 1:1 methanol/acetone fixative for 20 min at 4°C. Cells were washed thrice with 1xPBS containing 0.05% Tween 20 then blocked with a blocking buffer consisting of 2% bovine serum albumin (BSA) and 0.1% Tween 20 in 1×PBS for 1h. After washing 3 more times with wash buffer, SARS-CoV/SARS-CoV-2 nucleocapsid mAb (Sino Biological, Cat#40143-R001) diluted 1:5000 in 1×PBS containing 0.5% BSA and 0.1% Tween 20 was added to each well and incubated for 2h at 37°C. Detection antibody was removed by washing the plate 3 more times, then 1:2000 HRP-conjugated goat anti-rabbit polyclonal antibody (Dako, Denmark A/S, Cat#P0448) was added and the plate incubated at 37°C for 1h. Plates were washed thrice more, then TMB substrate was added (KPL, Cat#5120-0075) for 10 min. The reaction was stopped with 1N HCl. Absorbance was measured at 450 and 620 nm (reference wavelength) with an ELISA plate reader (Tecan Sunrise).

The average optical density (O.D.) at 450 and 620 nm were determined for virus control and cell control wells, and the neutralizing endpoint was determined by 50% specific signal calculation. The virus neutralizing endpoint titer of each serum was expressed as the reciprocal of the highest serum dilution with an OD value less than X which was calculated as follows:

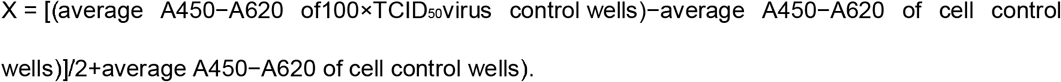

Sera which tested negative at 1:10 dilution were assigned a titer of <10. Sera were considered positive if neutralization titer was ≥20. Live SARS-CoV-2 viruses at passage 3 or 4 and Vero E6 cells at 20 maximum passages were employed. Activities with live viruses were carried out in a certified BSL-3 facility.

### Data and statistical analyses

The relationship between levels of (log-10 transformed) IgG titer, the immunogenic elicitor, serum collection date, age, and gender, was modelled using a linear model:

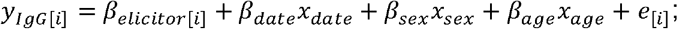

for immunogenic elicitor *i,y*_*IgG*[*i*]_ is (log-10 transformed) IgG titer; *x*_*date*_ is the serum collection date in days (from latest vaccine dose or from illness onset); *x*_*sex*_ indicates if the participant was a male; *x*_*age*_ is the age of the participant in years; *β*_*elicitor*[*i*]_ is the effect of the elicitor, while, *β*_*date*_, *β*_*sex*_, *β*_*age*_ and are the effects of the serum collection date, gender, and age, on the level of IgG titer respectively, and *e*_[*i*]_ is the residual error term. *e*_[*i*]_ are assumed to be independently and identically distributed (i.i.d.) 𝒩 (0, *σ*^2^). The *lm* function as implemented in the R library *stats* (v 4.0.4) was used to fit the model to the data. The level of IgG titer among the three cohort groups were compared using the Tukey method, adjusted for serum collection date, age, and sex, using the *emmeans* function as implemented in the R library *emmeans* (v 1.6.1).

By using the *lmer* function implemented in the R library *lme4*, a linear mixed model was fitted to the data to model the levels of NAb titers elicited by the three immunogenic elicitors against the four virus strains, accounting for the IgG titer, participant age and sex, as well as the variation among subjects:

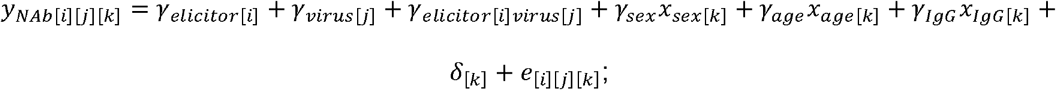

for immunogenic elicitor *i* virus *j* and subject *k,y*_*NAb*[*i*][*j*][*k*]_ is (log-10 transformed) NAb titer; *γ*_*elicitor*[*i*]_ is the effect of elicitor, *γ*_*virus*[*j*]_ is the effect of virus; *γ*_*elicitor*[*i*] *virus*[*j*]_ is the effect of the interaction between the elicitor and the virus; *x*_*sex* [*k*]_ indicates if the participant k was a male; *x*_*age*[*k*]_ is the age of the participant *k* in years; *x*_*IgG*[*k*]_ is (log-10 transformed) IgG titer of participant *k*; *γ*_*sex*_, *γ*_*age*_, *γ*_*IgG*_ and are the fixed effects of the gender, age, and IgG titer on the level of NAb titer, respectively; the varying coefficient *δ*_[*k*]_ accounts for the random effect raising form the fact that serum form a single participant k was used to measure NAb titers against the four virus strains under the study. *δ*_[*k*]_ is assumed to be normally distributed with mean zero and variant 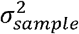, where 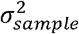 indicates the overall variation of the model intercepts among the participants, and similarly *e*_[*i*][*j*][*k*]_ ∼*i. i. d*. 𝒩(0, *σ*^2^). For each elicitor group, NAb titer values among virus groups were compared based on the model using the Tukey method with the *emmeans* function as implemented in the R library *emmeans* (v 1.6.1) (**Figure 2A**). The same analysis was performed to compare NAb titer values among cohort groups nested within virus strain (**Figure 2B**).

## Results

Our results demonstrated that a full 2-dose regimen of CoronaVac elicited levels of circulating SARS-CoV-2 S1-RBD-binding IgG (denoted ‘IgG titer’) that were comparable to levels detected in sera from both groups of naturally infected patients, adjusting for differences in participant age, gender, and serum collection date. Examination of the model revealed that age and gender were not significant predictors IgG titer (p = 0.414, and 0.682, respectively), but serum collection date significantly correlated with IgG titer (p = 2.02×10^−5^). 100% of participants in all cohorts were seropositive for IgG. (**Figure 1**).

**Figure 1.**
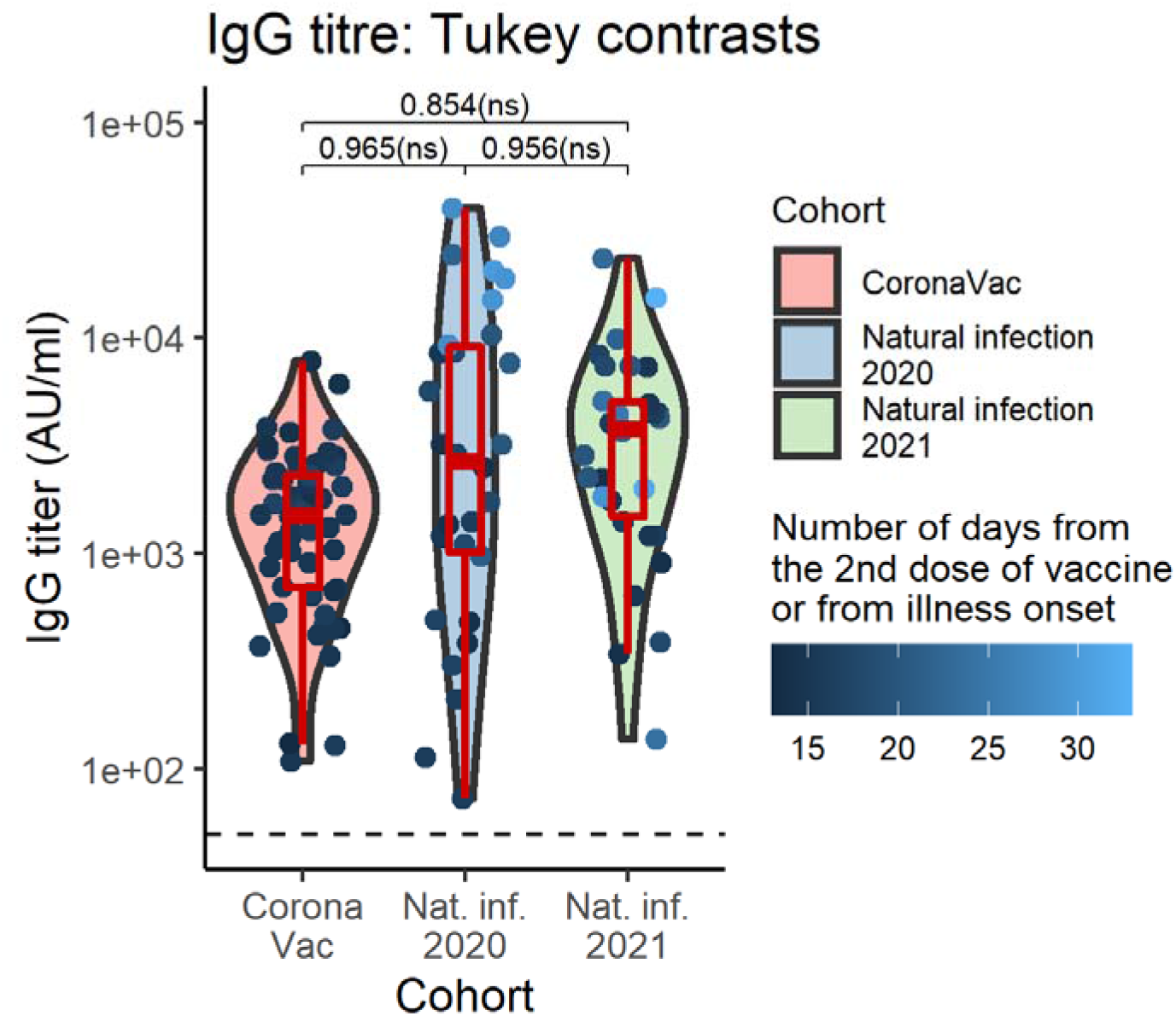
Violin plots of IgG titer by cohort group. Boxplots indicate medians and IQRs. The horizontal dotted line indicates the threshold for positive detection (50 AU/ml). There was no significant difference in SARS-CoV-2 S1-RBD-binding IgG levels between groups adjusting for patients’ sex, age, and serum collection date. Geometric mean IgG titer and 95%CI, computed at mean serum collection date (17.16 days), age (42.25 yr) averaged cross sexes: CoronaVac = 1741.28 (1240.48-2444.25) AU/ml; Natural infection 2020 = 1875.07 (1236.29-2843.89) AU/ml; Natural infection 2020 = 2034.54 (1340.47-3087.99) AU/ml. The data points are colored according to the serum collection date. The p values shown are Tukey-adjusted p-values.

We next assessed NAb-afforded protection against all 4 strains in our cohorts. Overall, the percentage of participants with quantifiable NAb titers above or equal to 20 units (the NAb positivity cut-off) was highest against the WT strain (99.17%), followed by Alpha (85.83%), and Beta (82.50%), and was lowest for the Delta strain (69.17%). This pattern was consistently observed for all cohorts and, notably, the percent of individuals with detectable NAbs were lowest in CoronaVac vaccinees compared to naturally infected groups (**Table 2**). After adjusting for participants’ age, sex, and IgG titer (fixed) effects as well as subject random effects, we indeed observed that, in all cohorts, there were statistically significant reductions in the mean NAb titers against all VOCs compared to WT. The NAb titers against the Alpha and Beta strains were not significantly different, and the NAb titers against the Delta strains were the lowest among all and were significantly different from the rest. **Figure 2A** summarizes the data, and the adjusted geometric mean NAb titers can be found in **Table 3**.

**Table 2.** Percentages of participants with NAb titers ≥20 (NAb positivity cut-off) against WT, Alpha, Beta and Delta strains within each cohort.

| NAb Titer ≥ 20 | WT | Alpha | Beta | Delta |
| --- | --- | --- | --- | --- |
| CoronaVac | 59/60 (98.33%) | 45/60 (75.00%) | 42/60 (70.00%) | 29/60 (48.33%) |
| Natural infection 2020 | 30/30 (100.00%) | 29/30 (96.67%) | 28/30 (93.33%) | 26/30 (86.67%) |
| Natural infection 2021 | 30/30 (100.00%) | 29/30 (96.67%) | 29/30 (96.67%) | 28/30 (93.33%) |
| Total | 119/120 (99.17%) | 103/120 (85.83%) | 99/120 (82.50%) | 83/120 (69.17%) |

**Table 3.** Predicted geometric mean NAb titer values, computed at mean age (42.25), geometric mean values of IgG titer (1824.71 AU/ml), and averaged across the sexes. The prediction assumed a linear mixed model, in which the effects of participants’ gender, age, (log-10 transformed) IgG titer, virus strain, and immunogenic elicitor on the observed values of (log-10 transformed) NAb titer were treated as fixed effects, and the effect of subject sampling was treated as a random effect. The model allowed the effects of virus strain and immunogenic elicitor to vary independently among each combination.

| Cohort | Virus strain | NAb titer |  |  |
| --- | --- | --- | --- | --- |
|  |  | Geometric mean | Lower 95% CI | Upper 95% CI |
| CoronaVac | WT | 774.48 | 607.22 | 987.82 |
|  | Alpha | 44.64 | 35 | 56.94 |
|  | Beta | 35.03 | 27.46 | 44.68 |
|  | Delta | 24.48 | 19.2 | 31.23 |
| Natural Infection 2020 | WT | 2329.83 | 1696.56 | 3199.49 |
|  | Alpha | 178.31 | 129.84 | 244.86 |
|  | Beta | 235.03 | 171.15 | 322.76 |
|  | Delta | 69.15 | 50.35 | 94.96 |
| Natural Infection 2021 | WT | 997.2 | 725.47 | 1370.72 |
|  | Alpha | 464.62 | 338.01 | 638.65 |
|  | Beta | 306.53 | 223 | 421.35 |
|  | Delta | 94.35 | 68.64 | 129.69 |

**Figure 2.**
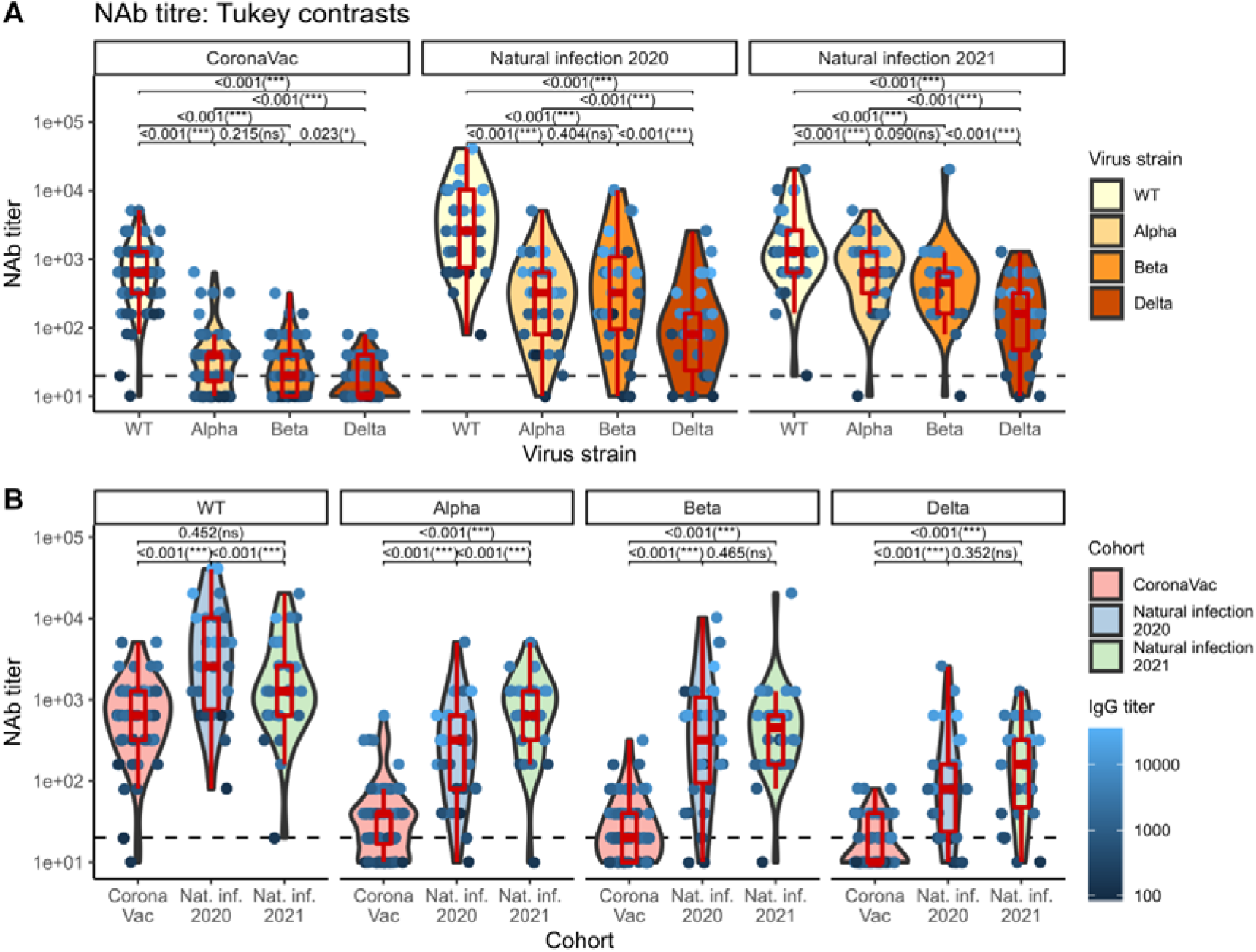
Violin plots of NAb titers grouped by virus strain nested within cohort groups (top), and by cohort group nested within virus strain (bottom). Boxplots indicate medians and IQRs. The horizontal dotted line indicates the threshold for positive detection (20 units). The data points are colored according to the IgG titer. The p-values shown are Tukey-adjusted p-values. IgG titer-, sex-, and age-adjusted geometric mean NAb titers can be found in **Table 3**.

We further found that WT was best neutralized by Natural Infection 2020 sera (NAb titer_geometric mean_ (NT_GM_) 2329.83 [95% CI 1696.56-3199.49]), 3.01 (1.84-4.92), and 2.34 (1.38-3.97) folds greater than CoronaVac and Natural Infection 2021 sera, respectively. The Alpha strain was best neutralized by Natural Infection 2021 sera (NT_GM_ 464.62 [95% CI 338.01-638.65]), 10.4 (6.35-17.1), and 2.61 (1.53-4.42) folds greater than CoronaVac and Natural Infection 2020 sera, respectively. These results are consistent with the predominant strains that circulated in Thailand in early/mid-2020 and mid-2021 at the time of sample collection for each respective cohort, which were the WT/D614G, and Alpha strains, respectively. Further, the Beta strain was neutralized equally well by Natural Infection 2020 and 2021 sera, with NAb titers (235.03 [95% CI 171.15-322.76] and 306.53 [95% CI 223-421.35], respectively) that were higher than that elicited by CoronaVac (NT_GM_ 35.03 [95% CI 27.46-44.68]), differing by 6.71 [4.10-11.0], and 8.75 [5.34-14.3] folds, respectively. Similarly, the Delta strain was neutralized equally well by Natural Infection 2020 and 2021 sera, but with markedly lower NAb titers than those obtained with the Beta strain (NT_GM_ 69.15 [95% CI 50.35-94.96] and NT_GM_ 94.35 [95% CI 68.64-129.69], respectively), and CoronaVac’s NAb titers were even lower still (NT_GM_ 24.48 [95% CI 19.2 -31.23]; being 2.82 [1.73-4.62], and 3.85 [2.35-6.32] folds lower in comparison, respectively), almost at the limit of detection (**Figure 2B**). Together, these results highlight the relatively low NAb titers elicited by CoronaVac compared to natural infection.

## Conclusions

Although NAb titers are not an exclusive immune correlate of protection, they are highly predictive of immune protection from symptomatic SARS-CoV-2 infection (3). Based on our data, although there was robust production of S1-RBD-binding IgG and 100% seropositivity, NAb-mediated protection was markedly reduced (and in many cases undetectable) against the 3 VOCs compared to WT in sera from all groups. NAb potency against Alpha and Beta were comparable in our CoronaVac vaccinee sera; this is inconsistent with a previous report that Beta shows more resistance to neutralization than Alpha in CoronaVac vaccinee sera collected 14 days after the second dose when tested using a pseudovirus neutralization assay (4). Worryingly, the Delta strain, which is the most transmissible, possibly among the most virulent of all VOCs identified to date (5), and is rapidly becoming a dominant strain in many countries, appears to be most refractory to neutralization. Lastly, our study highlights a low degree of neutralization-afforded protection mounted by CoronaVac when compared to natural infection. Further booster doses, heterologous or otherwise, beyond the conventional 2-dose regimen may be needed to maintain long-term anamnestic response. We also underscore a potential risk for reinfection in previously infected individuals, particularly with VOCs. Amid steady NAb decay over time (3) and the continued emergence of divergent SARS-CoV-2 strains, it is imperative to maintain effective mitigation strategies and to continue monitoring vaccine efficiency in areas with circulating VOCs.

## Data Availability

The data used to support the findings of this study are included within the article

## Ethics Statement

All methods were performed following standard protocols approved by the institutional review committee. The study protocol and human ethics were approved by the Human Research Ethics Committee, Faculty of Medicine Ramathibodi Hospital (COA. MURA2021/264) and the Ethics Committee of National Cancer Institute, Thailand (EC COA 019/2021).

## Acknowledgments

We would like to thank all the professionals in our COVID-19 research team and all the study participants. We also thank Dr. Man Van Minh Nguyen for helpful discussion regarding statistical analyses and Dr. Waradon Sungnak for helpful input.

